# Independently Carriage of *IL-1RN*2 Allele* Associated with Increased Risk of Gastric Cancer in The Sudanese Population

**DOI:** 10.1101/19013573

**Authors:** Abeer Babiker Idris, Amany Eltayib Ataelmanan, Sulafa Mohamed Eltaher, Einas Babiker Idris, Bashir M. Osman Arbab, Ahmed Ibn Idris, Mohamed Mansour, El-Amin Mohamed Ibrahim, Mohamed A. Hassan

## Abstract

**Background:** *Helicobacter pylori* is responsible for gastric cancer in approximately tens of millions of patients. Gastric cancer in Sudan represents one of the top causing death among cancers with about 686 cases per year and a 2.7 % mortality rate. *IL-1RN* VNTR polymorphism has been reported to increase the risk of gastric cancer.

**Objective:** The purpose of this study was to assess the association of the 86 bp VNTR polymorphism of *IL- 1RN* gene and the susceptibility to *H. pylori* infection and gastric cancer in the Sudanese population.

**Materials and methods:** Genomic DNA was extracted from 114 subjects. Of whom 60 had gastritis and duodenitis, 26 had a peptic ulcer, 16 had gastric cancer and 12 had normal gastroscopy findings. *H. pylori* infection was investigated by specific *16S rRNA*. And *IL-1RN* VNTR polymorphism at intron 2 was genotyped using the PCR method and direct sequencing for random samples.

**Results:** The positive *H. pylori* infection rate among participants was 47.37%. There is a lack of a significant difference in *IL- 1RN* genotype with *H. pylori* infection (p-value=1.0000). The *IL-1 RN L/L* genotype was significantly more frequent in a patient with benign disorders (gastritis or duodenitis or peptic ulcer), Odd=6.000 (95% CI =1.750-20.57, P=0.0056). While the heterozygote genotype 2/L was associated with an increased risk of gastric cancer with OR = 12.83 (95% CI = 1.261-130.6, P=0.0302).

**Conclusion:** Independently carriage of *IL-1RN *2* allele was associated with increased risk of gastric cancer in the Sudanese population. Notwithstanding the relatively small sample size of the study population, our findings show that the host genetic can be a useful tool for identifying high-risk individuals among dyspeptic patients; and also underscore the role played by host genetics in gastric carcinogenesis. To the best of our knowledge, this is the first study in Sudan concerning this issue.

## 1. Introduction

*Helicobacter pylori* infection is the most common human infection worldwide, approximately 50% of the world’s populations are infected and this is made human the main reservoir for *H. pylori*.^(1, 2)^ Although the high infection rates among the world population, most of them develop no clinical symptoms and continue their life with chronic gastritis.^(3-5)^ Unfortunately, still high percentages of the infected populations might develop severe gastric pathology. Approximately 17% of them will develop peptic ulcers and one-quarter of such patients even experience ulcer complications,^(6)^ while (approximately 1%) will progress to gastric cancers.^(6, 7)^ In terms of numbers, it is expected that several hundreds of millions will suffer from peptic ulceration and tens of millions might progress to gastric cancer.^(4)^

The presence of a pro-inflammatory response and the level of gastric acid secretion contribute significantly to the development of either peptic ulcer disease or atrophic gastritis.^(8, 9)^ IL-1 family has a pivotal role in the innate immunity and regulatory functions in the adaptive immune system.^(10)^ IL-1 (IL-1α and IL-1β), is the most influencing pro-inflammatory cytokine while *IL-1RN* is a naturally occurring anti-inflammatory cytokine.^(11-13)^ It is competitively bound to IL-1 receptors (IL-IR and IL-IIR), and thereby regulates the biological activity of IL-1α and IL-1β and modulates their potentially damaging effects.^(14-18)^ However, the therapeutic administration of IL-1ra, as a recombinant protein, has proven effective to prevent tissue damage^(10)^ and IL-1ra concentrations must be at least 100-fold higher than IL-1β concentrations to functionally block the biologic effects of IL-1β on target cells; hence, considerable production of IL-1ra is necessary for local tissues to stop the effects of IL-1β.^(19)^

The *IL-1* genes cluster spanning approximately 430 kb, which is located on the long arm of human chromosome 2q13-21, comprising *IL-1A, IL-1B, and IL-1RN. IL-1RN* (Location: 2q14.1, MIM:147679) encodes IL-1ra, the endogenous receptor antagonist of IL-1α and IL-1β, using the functional part of IL-1 thereby modulates their potentially damaging effects on target cell.^(15, 20, 21)^ The *IL-1RN* gene has been linked with a broad range of chronic inflammatory diseases and enhanced IL-1β secretion.^(11, 22-26)^ The imbalance between IL-1 and IL-1ra has a crucial impact in developing inflammatory disorders in local tissues taking part in numerous illnesses, including infectious diseases, through overproduction of IL-1α, β and/or underproduction of IL-1ra.^(27-29)^

Polymorphisms in *IL-1* and *IL-1RN* have been reported to be related to variations in the production levels of IL-1 (IL-1B and IL-1A) and IL-1ra.^(13, 30, 31)^ The most intensively studied IL-1RN polymorphism connected to susceptibility to *H. pylori* infection and gastric cancer outcome is a penta-allelic 86-bp *VNTR* (variable number of tandem repeats) polymorphism that is located in intron 2.^(12, 32-34)^ The most typical variants contains four repeats named allele *IL1RN*1* and variants with 2 repeats (allele *IL1RN*2*). While other alleles, 3 repeats (allele *IL1RN*4*), 5 repeats (*allele IL1RN*3*) and 6 repeats (*allele IL1RN*5*),^(35)^ occur at a combined frequency of <5%.^(14)^ Vamvakopoulus *et. al*., in 2000, revealed the sixth variant of this polymorphism containing only one repeat (allele *IL1RN*0*).^(36)^ Studies from different ethnic groups have reported that in individuals infected with *H. pylori, IL-1RN*2* allele is associated with increased production of IL-1β which leading to severe and sustained inflammation, gastric atrophy, hypochlorhydria, and ultimately to the development of gastric cancer.^(22, 31, 32, 37-39)^ Contrariwise, Asian studies have been less conclusive on the relation between *IL-1RN* polymorphism, *H. pylori* infection and gastric cancer.^(40, 41)^ It seems that there is an ethnic variability with regard to *IL-1RN* polymorphisms frequency and their association with *H. pylori* infection and gastric cancer. To the best of our knowledge, there have been no previous studies in Sudan have investigated the role of the *IL-1RN* polymorphisms in *H. pylori* infection and gastric cancer. Therefore, the purpose of this study was to assess the association of the 86 bp VNTR polymorphism of *IL-1RN* gene and the susceptibility to *H. pylori* infection and gastric cancer in the Sudanese population.

## 2. Materials and Methods

### 2.1 Ethical consideration

This study was approved by the Khartoum ministry of the health research department, University of Khartoum and Research Ethics Committees of hospitals.

### 2.2 Study design and Study sittings

The study was conducted in major public and private hospitals in mainly Khartoum state from June 2018 to January 2019. These hospitals include Ibin Sina specialized hospital which is a public hospital. And three private hospitals which include Modern Medical Centre, Al-Shorta hospital and Al Faisal Specialized Hospital. The distribution of participants’ samples from hospitals in percentage is given in Figure1. All sample processes were carried out in the Molecular biological lab at the Faculty of Medical Laboratory Sciences at the University of Khartoum.

**Figure 1.**
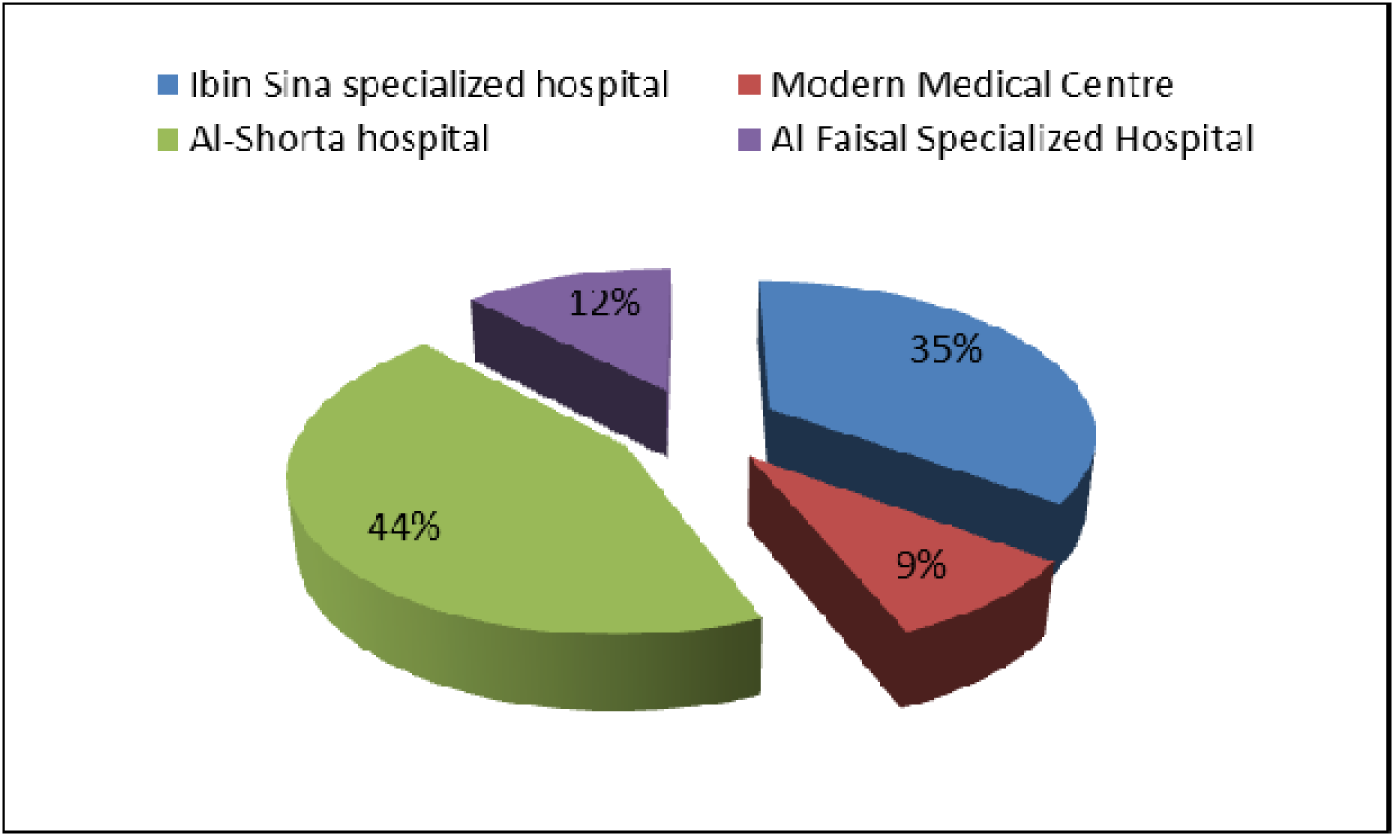
Distribution of participants among hospitals

### 2.3 Study population

The study population composed of 114 subjects consisted of 54 *H. pylori* positive patients (33 male) of whom 29 had gastritis or/and duodenitis, 16 had a peptic ulcer, and 4 had gastric cancer, and 60 were *H. pylori* negative subjects (34 males) who regarded as controls, see Table 1. The subjects were recruited from the Endoscopy unit of public and private hospitals in Khartoum state. The diagnosis of gastroduodenal diseases was based on the assessment of an experienced gastroenterologist and the investigation of gastric cancer was confirmed by histopathology. The distribution of participants according to endoscopy series and clinical symptoms were presented in Tables 2 and 3, and Figure 2.

**Table 1.** Demographic characteristic of participants

| Variables |  | Total<br>n=114 | *H. pylori* (+ve)<br>n=54 | *H. pylori* (-ve)<br>n=60 | P-value |
| --- | --- | --- | --- | --- | --- |
| **Age years ± Std. Deviation (range)** | | 44.91±17.38<br>(15-89) | 44.67±16.79<br>(15-85) | 45.13±18.03<br>(17-89) | 0.9864 |
| **Gender** | Male | 67 (58.77%) | 33 (49.25%) | 34 (50.75%) | 0.7046 |
|  | Female | 47 (41.23%) | 21 (44.68%) | 26 (55.32%) |  |
| **Residence** | Urban | 49 (42.98%) | 19 (38.78%) | 30 (61.22%) | 0.132 |
|  | Rural | 65 (57.02%) | 35 (53.85%) | 30 (46.15%) |  |
| **Hospital** | Public | 41 (35.96%) | 13 (31.71%) | 28 (68.29%) | **0.019** |
|  | Private | 73 (64.04%) | 41 (56.16%) | 32 (43.84%) |  |

**Table 2.** Distribution of participants according to endoscopy series and status of *H. pylori* infection

| Endoscopy results | No.(%) | *H. pylori*<br>(+ve) | *H. pylori*<br>(-ve) | P-value |
| --- | --- | --- | --- | --- |
| **Normal gastric findings** | 12 (10.53%) | 5 (41.67%) | 7 (58.33%) | 0.4319 |
| **Gastritis** | 55 (48.25) | 27 (50%) | 28 (46.67%) |  |
| **Duodenitis** | 5 (4.39%) | 2 (3.70%) | 3 (5%) |  |
| **Gastric ulcer** | 17 (14.91%) | 11 (20.37%) | 6 (10%) |  |
| **Duodenal ulcer** | 5 (4.39%) | 3 (5.56%) | 2 (3.33%) |  |
| **Gastroduodenal ulcers** | 4 (3.51%) | 2 (3.70%) | 2 (3.33%) |  |
| **Carcinoma** | 16 (14.03%) | 4 (7.4%) | 12 (20%) |  |
| **Total** | 114 (100%) | 54 (100%) | 60 (100%) |  |

**Table 3.**
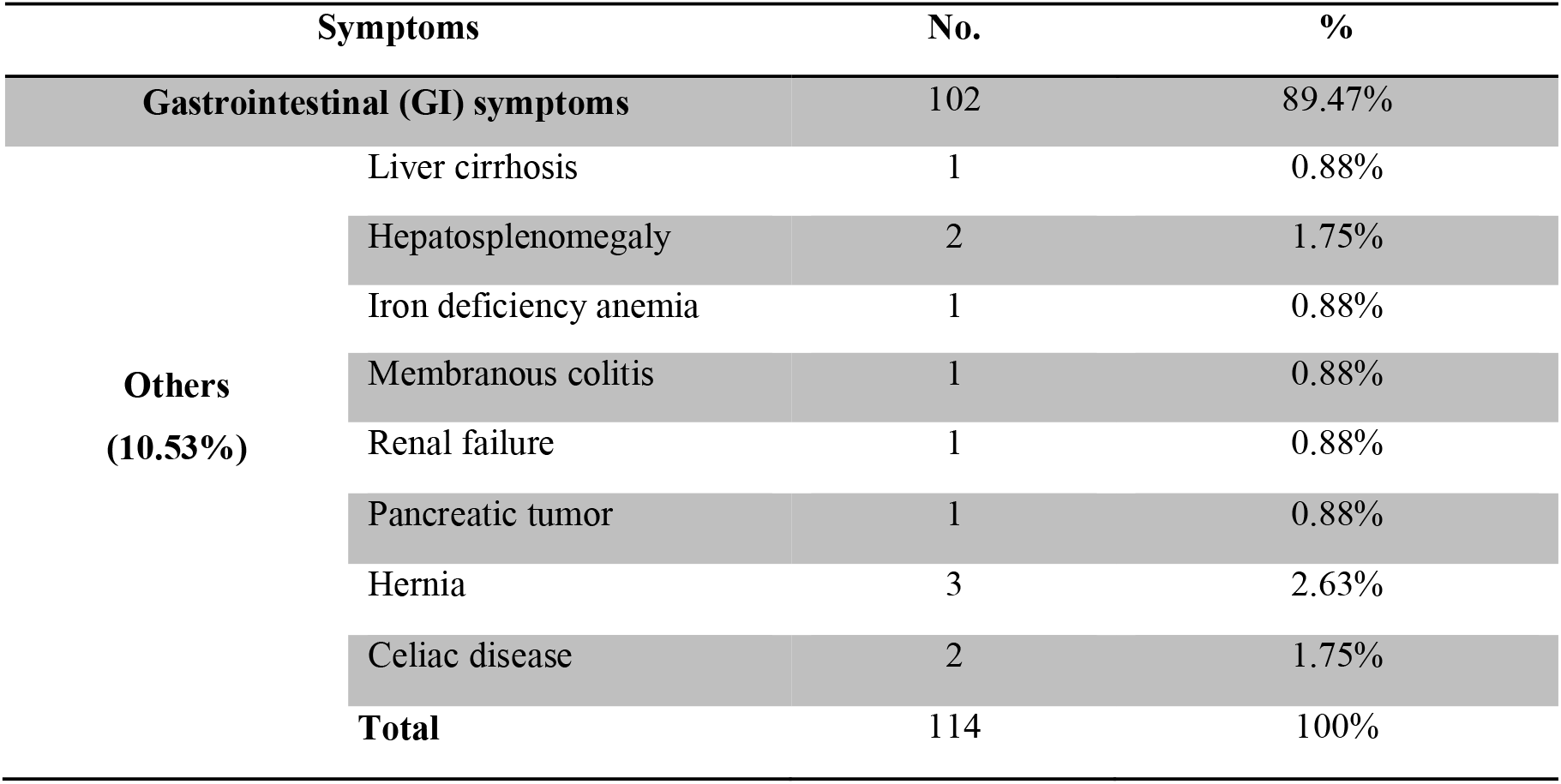
Distribution of participants according to clinical symptoms

**Figure 2.**
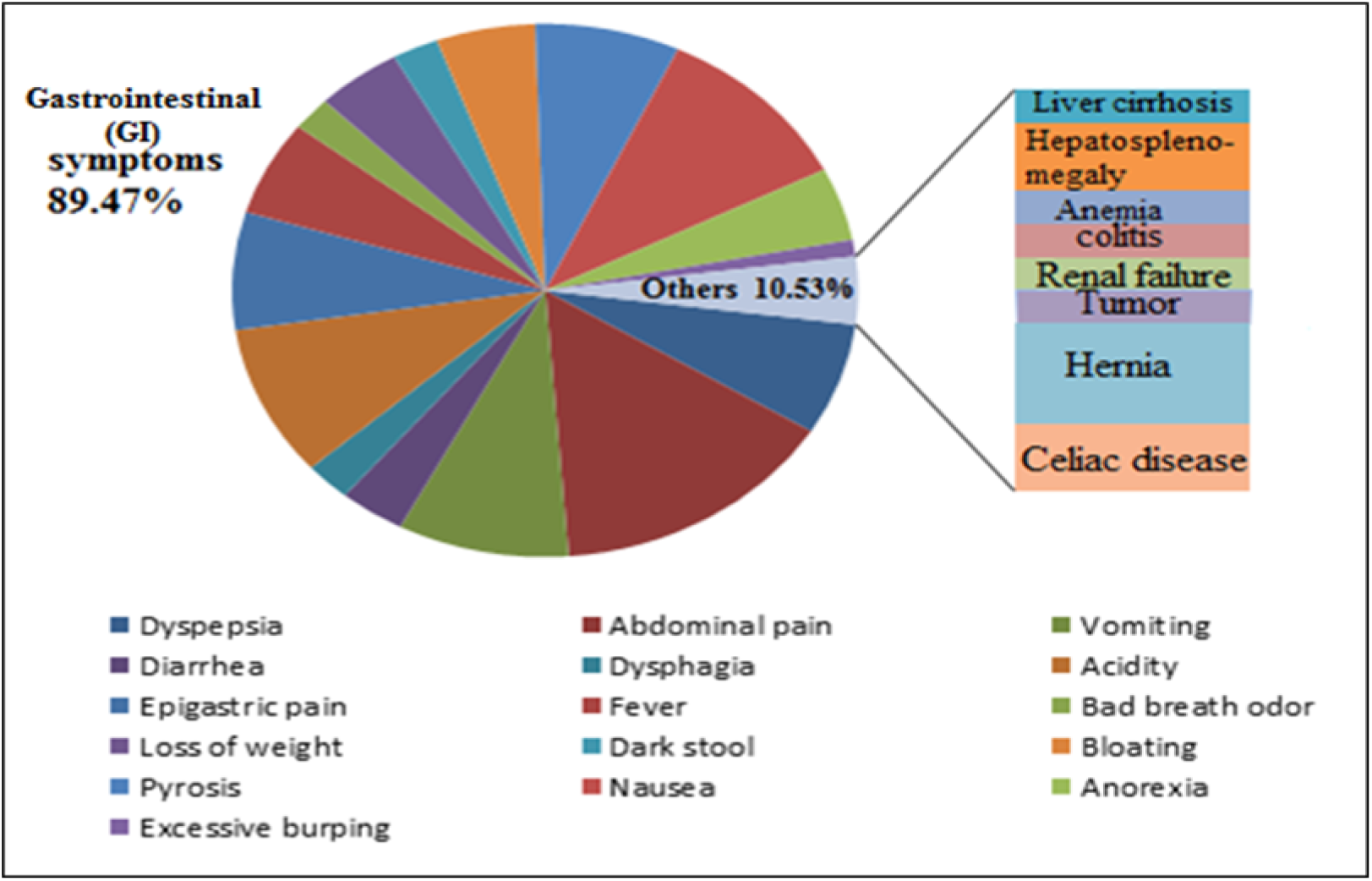
Shows clinical symptoms of participants

Each participant was interviewed using a structured questionnaire to seek information on residence, gastroenterological symptoms, and other variables. All interviews were conducted in the hospital before enrolling the participants. The selection criteria included the Sudanese population from both sexes, no antibiotic or NSAIDS uses. All the participants were informed with the objectives and purposes of the study and the written informed consents were taken.

### 2.4 Sample collection

A total of 114 gastric biopsies were collected in 1.5µl Eppendorf tubes with 400µl phosphate buffer saline (PBS). The biopsies were labeled and transported immediately to the molecular biological Lab then stored at −20°C until complete the collection of samples.

### 2.5 DNA extraction

The DNA extraction was done by using the innuPREP DNA Mini Kit (analytikjena AG, Germany) and performed according to the instructions of the manufacturing company. In a brief, the extraction procedure is based on a new kind of chemistry. First, lysis of biopsies which was done by adding 400µl of TLS and 25µl of proteinase K; and vortex for 5sec then incubation at 50°C until complete lysis occurred. Then, Centrifuging at 100rpm for 1min and transferring the supernatant to a new 1.5ml Eppendorf. Secondly, efficient binding of the DNA on a Spin Filter surface by adding 400µl of TBS and vortex for 15sec then adding the sample to the Spin Filter and centrifuge at 12000rpm for 2min. thirdly, washing the bound DNA by adding 500µl HS and centrifuge at 12000rpm for 1min then adding 750µl MS and centrifuge at 12000rpm for 1min. Finally, eluting of the DNA by adding the Spin Filter to an Elution Tube then adding 200µl of the Elution Buffer and incubation for 1min at room temperature then centrifuged at 8000 for 1min. The extracted DNAs were stored at −20°C until further processing.

### 2.6 *H. pylori* 16S rRNA specific gene amplification

Extracted DNA was amplified for the specific 16S rRNA gene to investigate the infection of *H. pylori*. (primers: F:5’-GCGCAATCAGCGTCAGGTAATG-3’) (R:5’-GCTAAGAGAGCAGCCTATGTCC-3’). ^(42)^

PCR amplification was performed with Maxime PCR PreMix Kit (i-Taq) (iNtRON BIOTECHNOLOGY, Seongnam, Korea) and a PCR thermocycler (SensoQuest, Germany). The reaction mixtures used for PCR contained 2.5U of i-Taq TM DNA polymerase (5U/µl), 2.5mM of each deoxynucleoside tri-phosphates (dNTPs), 1X of PCR reaction buffer (10X),1X of gel loading buffer and 1µl of DNA template. The temperature cycle for the PCR was an initial step of 3min at 94°C, followed by denaturation for 30sec at 94°C, annealing for 30sec at 53°C and primer extension for 45sec at 72°C. After the 40th cycle, the final extension step was prolonged for 5min to complete the synthesis of strands.

PCR products were detected by gel electrophoresis. 3µl of each PCR products was loaded onto 2% agarose gels stained with 3µl ethidium bromide (10mg/ml) and subjected to electrophoresis in 1x Tris EDTA Buffer (TEB buffer) (89mM of Tris base, 89mM Boric acid and 2mM EDTA dissolved in 1Litter H2O) for 30 min at 120V and 50mA. The gel was visualized under UV light illumination. A 100MW DNA ladder (iNtRON BIOTECHNOLOGY, Seongnam, Korea) was included in each gel as a molecular size standard. The amplified products for the specific 16SrRNA gene is 522bp.

### 2.7 Detection of the IL-1RN 86bp VNTR at intron2 using conventional PCR

The IL-1RN 86bp VNTR was analyzes as previously described. ^(43)^ Briefly, oligonucleotides primers for PCR amplification were 5’-CTCAGCAACACTCCTAT-3’ and 5’-TCCTGGTCTGCAGGTAA-‘3. The amplification conditions were 94°C for 1min, 35 cycles of 94°C for 1min, 60°C for 1min, 72°C for 1min, and a final extension at 72°C for 7 minutes. The PCR products were visualized by electrophoresis on a 2% agarose gel stained with ethidium bromide. A 1-kilobase DNA ladder was used to standardize alleles’ sizes and code conventionally as follows: allele 1 (A1)=410bp (4 repeats), allele 2 (A2)=240bp (2 repeats), allele 3 (A3)=500bp (5 repeats), allele 4 (A4)=325bp (3 repeats), and allele 5 (A5)=595bp (6 repeats). For statistical analysis purposes and in accordance with a previous report conducted by Machado *et al*., these alleles are classified into the short (allele *2=*2) and long alleles (alleles *1, *3, 4, and 5=L). The genotyping patterns will classify as L/L, L/*2, and *2/*2.^(38, 44)^

### 3.8 Sequencing and sequence analysis

DNA purification and Sanger dideoxy sequencing were performed commercially by Macrogen Inc, Korea. For each sequence, two chromatograms (forward and reverse), were visualized and analyzed by using the Finch TV program version 1.4.0.^(45)^ The Basic Local Alignment Search Tool (BLAST; https://blast.ncbi.nlm.nih.gov/) was used to look for and assess nucleotide sequence similarities; also to check the quality of our PCR procedure for detecting of VTRP *IL-1RN*.^(46)^

### 2.9 Statistical analysis

Hardy-Weinberg equilibrium of genotype distributions was assessed by χ^2^ test or *Fisher*’s test. Descriptive statistical analysis was performed to calculate proportions, percentages, means, and standard deviations. According to the prevalence of *H. pylori* infection, differences in frequency distribution by age were assessed by the Mann-Whitney test. While differences in frequency distribution among the categorical demographic characteristics of the study population were examined with χ^2^ test or *Fisher’*s test. Associations of I*L-1RN* genotypes with *H. pylori* infection and endoscopic series were assessed by odd ratios (ORs) and 95% confidence intervals (CIs) using unconditional logistic regression analysis. Differences were considered to indicate statistical significance if P<0.05. Data were analyzed using the Statistical Package for Social Sciences (SPSS version 19).

## 3. Result

The positive *H. pylori* infection rate among participants was 47.37%. No significant differences were noted for *H. pylori* infection rates by age, gender, and residence. A significant association was found with the type of hospitals (p=0.019) where participants from private hospitals involved a high percentage of infection.

Fifty-four patients and fifty-seven of sixty uninfected subjects were successfully typed for the *IL-1RN* polymorphism. In this study, six *IL-1RN* genotypes (1/1, 1/2, 2/2, 1/3, 2/3, and 1/4) were included while allele 5 (6 repeats) and allele 6 (0 repeats) of IL-1ra and homozygotes of the rare alleles (A3 and A4) were not detected in both groups of participants (infected and uninfected), see Figure 3. The frequency of *IL-1RN* alleles is presented in Table4. *IL-1RN* allelic distributions did not deviate significantly from those expected in Hardy-Weinberg equilibrium for control subjects (for uninfected subjects p=0.5923; and for subjects with normal gastroscopy series p=0.5983).

**Table 4.** Four alleles of the *IL-1RN* 86bp VNTR at intron2 observed among participants

| Allele | Frequency | Size (bp) | Number of repeats |
| --- | --- | --- | --- |
| A1 | 0.893 | 410 | 4 repeats |
| A2 | 0.094 | 240 | 2 repeats |
| A3 | 0.009 | 500 | 5 repeats |
| A4 | 0.004 | 325 | 3 repeats |

**Figure 3.**
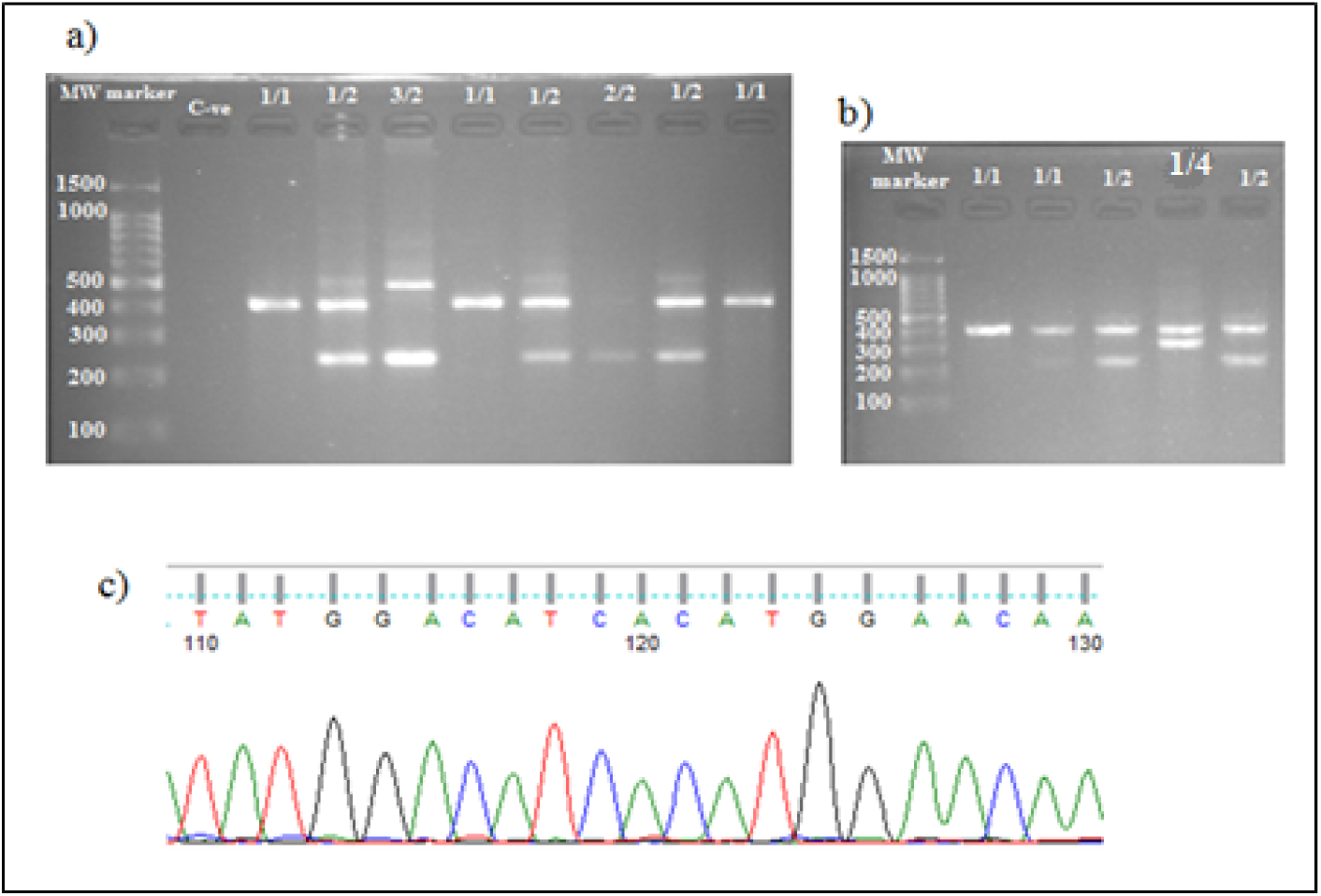
3a) and 3b) Illustrate PCR products analyzed on a 2% agarose gel stained with ethidium bromide. Four different alleles can be seen at 240bp, 325bp, 410bp and 500bp representing 2, 3, 4 and 5 copies of the 86-bp sequence, respectively. 3c) shows the sequencing result of the chromatogram using Finch TV software. The 4 nucleotide sequences of the *IL-1RN* containing 86bp VNTR polymorphism were deposited in the GenBank database under the following accession numbers: from MN641824 to MN641829

The present study found a lack of a significant difference in *IL-1RN* genotype with *H. pylori* infection (p-value=1.0000), as shown in Table 5. The homozygote allele 2* (2/2 repeats) and the heterozygote allele 3* and allele 4* were found only in patients infected with *H. pylori*, one patient for each.

**Table 5.** Allele and genotype frequencies of *IL-1RN* VNTR polymorphism among *H. pylori* infected and uninfected subjects, and their contributions to *H. pylori* infection

|  | *H. pylori* (+ve)<br>n=54 | *H. pylori* (-ve)<br>n=57* | OR (95% CI) |
| --- | --- | --- | --- |
| Allele frequency |  |  |  |
| *IL-1RN* *L | 96 (88.89%) | 103 (90.35%) | 0.8544 (0.3600-2.028) |
| *IL-1RN* *2 | 12 (11.11%) | 11 (9.65%) |  |
| P-value | 0.8267 |  |  |
| Genotype frequency |  |  |  |
| L/L | 43 (79.63%) | 46 (80.70%) | 1 |
| 2/L | 10 (18.52%) | 11 (19.30%) | 1.028 (0.3968-2.665) |
| 2/2 | 1 (1.85%) | 0 | 0.3118 (0.01236-7.867) |
| 2/L + 2/2 | 11 (20.37%) | 11 (19.30%) | 0.9348 (0.3675-2.378) |
| L/L * L/2+2/2 | 1.0000 |  |  |
| P-value |  |  |  |

\*3 Samples are missing

A profile of *IL-1RN* polymorphism in relation to gastric pathologies reveals that the *IL-1RN*L* allele was significantly associated with gastric pathologies risk if all patients were included (P=0.0261), see Table 6. The *IL-1 RN L/L* genotype was significantly more frequent in a patient with benign disorders (gastritis or duodenitis or peptic ulcer), Odd=6.000 (95% CI =1.750-20.57, P=0.0056), as shown in Table 7. Moreover, there is a significant association between gastric cancer and the carriage of *IL-1RN *2* allele (P=0.0504), see Table 6. The homozygous *ILRN 2/2* genotype was found in only one patient with antral gastritis while the heterozygote genotype 2/L was associated with an increased risk of gastric cancer with OR = 12.83 (95% CI = 1.261-130.6, P=0.0302). The risk for *IL-1RN*2* allele carriers increased by 15.75 (95% CI=1.448-171.3, P = 0.0235) as shown in Table 8. The gastric cancer risk was more evident for *IL-1RN* 2/L with OR=18.00 (95% CI =1.631-198.6, P = 0.0183).(data not shown). However, the calculation of the odds ratio of *ILRN 2/L* did not reach a statistical significance with gastritis/duodenitis or peptic ulcer, see Table 6 for more illustration.

**Table 6.** Allele and genotype frequencies of *IL-1RN* VNTR polymorphism in different gastric pathologies

| Gastric pathologies |  |  |  |  |  |  |  |
| --- | --- | --- | --- | --- | --- | --- | --- |
|  | Normal<br>finding<br>n=12 | Gastritis<br>and / or<br>Duodenitis<br>n=60 | OR<br>(95% CI) | Peptic<br>ulcer<br>n=26 | OR<br>(95% CI) | Gastric<br>cancer<br>n=13 | OR<br>(95% CI) |
| Allele frequency |  |  |  |  |  |  |  |
| *IL-1RN* *L | 23<br>(95.8%) | 109<br>(90.83%) | 2.321<br>(0.2852-18.89) | 48<br>(92.3%) | 1.917<br>(0.2025-18.14) | 19<br>(73.1%) | 8.474<br>(0.9558-75.12) |
| *IL-1RN* *2 | 1<br>(4.16%) | 11<br>(9.16%) |  | 4<br>(7.69%) |  | 7<br>(26.9%) |  |
| P value | 0.6908 |  |  | 1.0000 |  | 0.0504 |  |
|  | 0.0261 |  |  |  |  |  |  |
| Genotype frequency |  |  |  |  |  |  |  |
| L/L | 11<br>(91.67%) | 50<br>(83.3%) | 1 | 22<br>(84.6%) | 1 | 6<br>(46.2%) | 1 |
| L/2 | 1<br>(8.3%) | 9<br>(15%) | 1.980<br>(0.2267-17.29) | 4<br>(15.4%) | 2.000<br>(0.1989-20.11) | 7<br>(53.8%) | 12.83<br>(1.261-130.6) |
| 2/2 | 0 | 1<br>(1.67%) | 0.6832<br>(0.0261-17.88) | 0 | - | 0 | - |
| L/2+2/2 | 1<br>(8.3%) | 10<br>(16.67%) | 2.200<br>(0.2544-19.03) | 4<br>(15.4%) | 2.000<br>(0.1989-20.11) | 7<br>(53.8%) | 12.83<br>(1.261-130.6) |
| L/L * L/2+2/2 | 0.6772 |  |  | 1.0000 |  | 0.0302 |  |
| P value | 0.0109 |  |  |  |  |  |  |

**Table 7.**
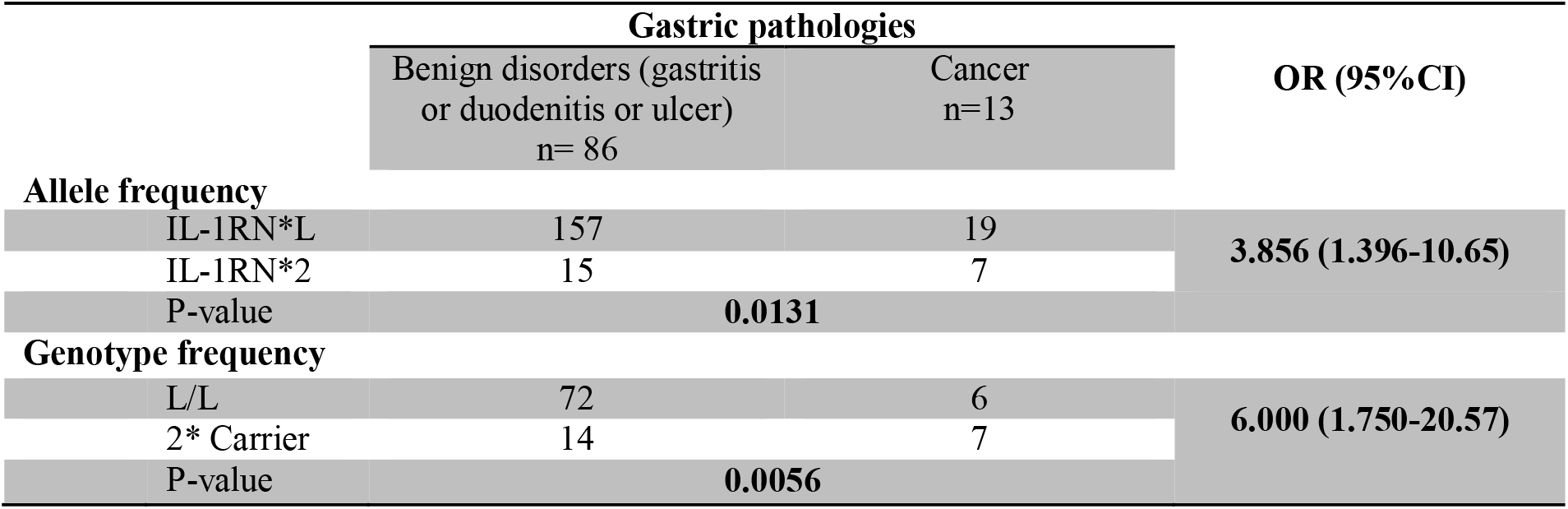
*IL-1RN* VNTR polymorphism and gastric cancer association risk in gastric cancer patients and subjects with benign gastric disorders

| Gastric pathologies |  |  |  |
| --- | --- | --- | --- |
|  | Benign disorders (gastritis<br>or duodenitis or ulcer)<br>n= 86 | Cancer<br>n=13 | OR (95%CI) |
| Allele frequency |  |  |  |
| IL-1RN*L | 157 | 19 | 3.856 (1.396-10.65) |
| IL-1RN*2 | 15 | 7 |  |
| P-value | 0.0131 |  |  |
| Genotype frequency |  |  |  |
| L/L | 72 | 6 | 6.000 (1.750-20.57) |
| 2* Carrier | 14 | 7 |  |
| P-value | 0.0056 |  |  |

**Table 8.**
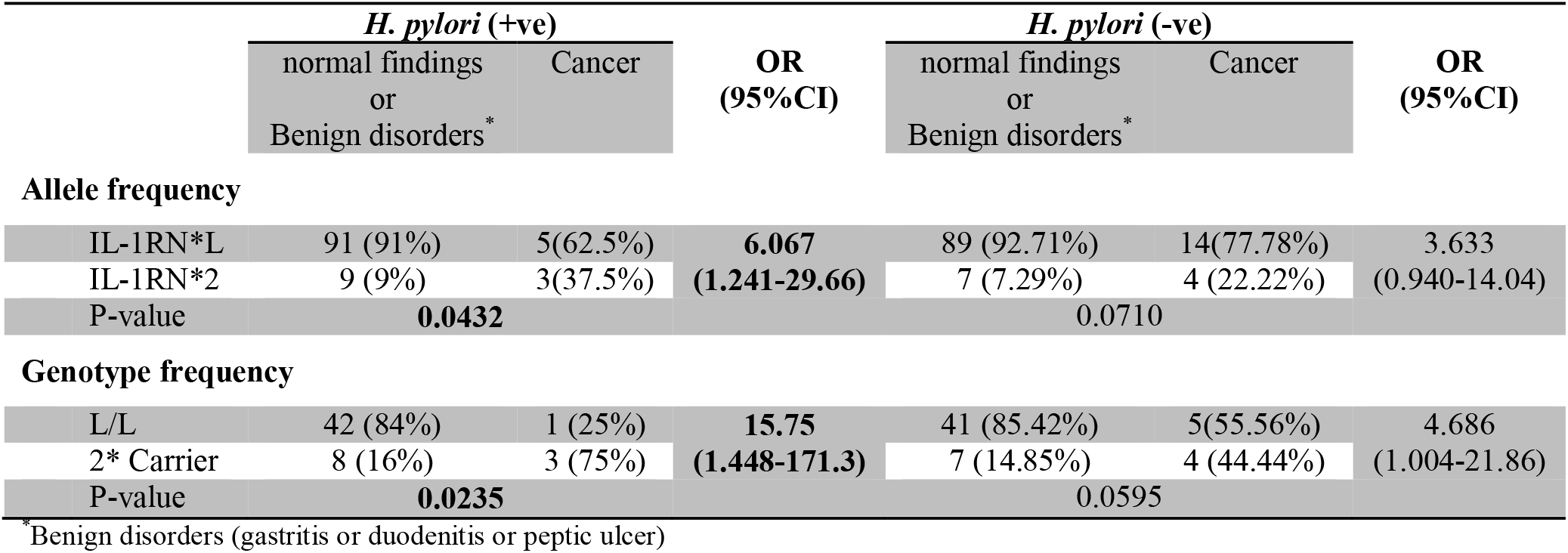
Gastric cancer risk association with *IL-1RN* polymorphism in 13 gastric cancer patients and 89 control subjects with normal findings or benign disorders according to *H. pylori* status

|  | *H. pylori* (+ve) |  | OR<br>(95%CI) | *H. pylori* (-ve) |  | OR<br>(95%CI) |
| --- | --- | --- | --- | --- | --- | --- |
|  | normal findings<br>or<br>Benign disorders * | Cancer |  | normal findings<br>or<br>Benign disorders * | Cancer |  |
| Allele frequency |  |  |  |  |  |  |
| IL-1RN*L | 91 (91%) | 5(62.5%) | 6.067 | 89 (92.71%) | 14(77.78%) | 3.633 |
| IL-1RN*2 | 9 (9%) | 3(37.5%) | (1.241-29.66) | 7 (7.29%) | 4 (22.22%) | (0.940-14.04) |
| P-value | 0.0432 |  |  | 0.0710 |  |  |
| Genotype frequency |  |  |  |  |  |  |
| L/L | 42 (84%) | 1 (25%) | 15.75 | 41 (85.42%) | 5(55.56%) | 4.686 |
| 2* Carrier | 8 (16%) | 3 (75%) | (1.448-171.3) | 7 (14.85%) | 4 (44.44%) | (1.004-21.86) |
| P-value | 0.0235 |  |  | 0.0595 |  |  |

\*Benign disorders (gastritis or duodenitis or peptic ulcer)

## 4. Discussion

In this study, the analysis of *IL-1RN* 86bp VNTR polymorphism at intron 2 showed a significant association between gastric cancer and independently carriage of *IL-1RN *2* allele (P=0.0504) in the Sudanese population. This finding is in agreement with other studies from Portugal, Italy, and the UK which also found that the presence of *IL-1RN *2* increased the risk of gastric cancer.^(31, 38, 47, 48)^ Since 2000 when Omer *et al*., reported the first significant association between gastric cancer risk and polymorphism in the genes *IL-1B* and *IL-1RN* among Scottish and Polish patients,^(31)^ a lot of studies from different ethnic groups and populations have pointed out this relation as well.^(32, 49, 50)^ Unfortunately, to the best of our knowledge, no data regarding these polymorphisms are available in Sudan where gastric cancer, according to GLOBOCAN 2018, represents one of the top causing death among cancers with about 686 cases per year and 2.7 % mortality rate.^(51)^ However, most of the studies have observed this association with an *IL-1RN* 2/2 homozygous genotype^(31, 38, 48)^ but we found this association with a heterozygous genotype (*IL-1RN* 2/L) with OR=12.83 (95% CI = 1.261-130.6, P = 0.0302) which is consistent with other studies conducted in Italy,^(47, 52)^ Brazil,^(53)^ Taiwan,^(54)^ China,^(55)^ Turkey^(39)^ and Arabic population (Omani and Algerian)^(56, 57)^ that found the same association of gastric cancer with a *IL-1RN* 2/L heterozygous genotype.

Moreover, the risk for *IL-1RN *2* carriers increased by15.75 (95% CI=1.448-171.3, P = 0.0235) in *H. pylori* infected patients. Similarly, a significantly increased risk of gastric cancer with *IL-1RN *2* carriers who infected with *H. pylori* was noticed with a moderate increase among Hispanics, but not among the Asian population. ^(23, 58-60)^ In 2005, Ruzzo *et al*. studied the potential etiologic role of *IL-1RN* VNTR polymorphism in *H. pylori*-negative, Italian gastric cancer patients. And he found that the *IL-1RN\**2 allele may contribute to intestinal gastric cancer risk in the absence of concomitant *H. pylori* infection which suggests that the *IL-1RN\**2 allele could contribute to disease risk independently.^(47)^ However, other dietary environmental mutagens and carcinogens may interact with the pro-inflammatory cytokines on the gastric mucosa then induce gastric cancer even in the absence of *H. pylori* infection.^(61, 62)^ N-nitroso compounds and polycyclic aromatic hydrocarbons have been found to increase IL-1β production levels. ^(63-65)^ Moreover, *IL-1RN* and *IL-1B* have been reported to independently enhance cell proliferation and tumorigenesis. ^(66, 67)^

The variation in *IL-1RN* gastric cancer predisposition risk between studies could be due to the different genetic backgrounds in different ethnic groups or due in part to the variation in the frequency of the *IL-1RN* 2/2 homozygous genotype in the study populations. In our study, only one patient had *IL-1RN* 2/2 homozygous genotype and actually, a potential limitation of our investigation is the relatively small sample size. A low prevalence of *IL-1RN* allele 2 has observed previously in African-American^(68)^ and black African populations. ^(69, 70)^ The frequency of the *IL-1RN* *2 allele in the Sudanese study population was 9.4% which is low compared with 26% and 15% in Caucasian and Omani populations, respectively but high in comparison with the Asian population for whom frequencies of around 6% are reported.^(31, 41, 52, 56)^

In the African population, data are scarce regarding the effects of *IL-1RN* VNTR polymorphism on gastric pathology. In this study, we found a significantly prevalent of *IL-1 RN L/L* genotype in patients with benign disorders (gastritis or duodenitis or peptic ulcer), Odd=6.000 (95% CI =1.750-20.57, P=0.0056). This finding is in concordance partially with the result reported by Kimang’ a *et al*. in an African population in 2012 that the *IL-1RN* L/L were significantly more frequent in *H. pylori* associated gastric pathologies (gastritis, peptic ulcer disease, and gastrointestinal reflux disease). ^(70)^

With regard to the susceptibility to *H. pylori* infection, we found a lack of association between *IL-1RN* 86bp VNTR polymorphism and susceptibility to *H. pylori* infection. This negative finding is similar to those reported in Japanese and UK populations. ^(44, 71)^ The true nature of the relationship between the polymorphisms of pro-inflammatory cytokines, susceptibility to *H. pylori* infection and development of gastric cancer is likely to be extremely complex and still unclear. Although the high prevalence of *H. pylori* infection (about 50% of the world’s population), only 15% of these individuals go on to develop clinically significant diseases.^(72)^ The accepted model for the variation in *H. pylori*-associated diseases relates to the amount of stomach acid secretion, the site of *H. pylori* colonization and the distribution of the infection; i.e., antral gastritis favors duodenal ulcer formation whereas corpus-predominant gastritis favors the development of a gastric ulcer and sometimes progressing to metaplasia and adenocarcinoma. The sever impaired acid secretion that typically results from diffuse gastritis allows *H. pylori* to colonize the corpus. ^(73, 74)^ However, *IL-1B* and *IL-1RN* polymorphisms increase IL-1β production which suppresses gastric acid secretion and therefore potentially influences *H. pylori* distribution within the stomach and predisposes individuals to gastric cancer. ^(31, 37)^ These facts together lead us to suggest that the genetic association of *IL-1RN* polymorphisms with gastric cancer is a result of the complex interaction between the host inflammatory response and *H. pylori* but not primarily related to increased susceptibility to *H. pylori* infection. Moreover, a number of studies conducted in different ethnic groups showed an association between *IL-1RN* polymorphisms and *H. pylori*-mediated diseases (gastritis, duodenitis, ulcer, and cancer), see Table 9 for more illustration.

**Table 9.** Summary of published studies on the association between the susceptibility to *H. pylori* infection, gastroduodenal diseases and *IL-1RN* 86bp VNTR polymorphism in different ethnic groups

| Author | Year | Country | Ethnicity | Screening method | Inference | Reference |
| --- | --- | --- | --- | --- | --- | --- |
| **Abeer** | 2019 | Sudan | Sudanese | PCR and direct sequencing | *IL1-RN* 2/L increases risk of gastric cancer | This study |
| **Lu W** | 2005 | China | Chinese | PCR-based denaturing high-performance liquid chromatography analysis and direct DNA sequencing | No association between the susceptibility to *H. pylori* infection, gastric cancer and *IL-1RN* polymorphism | (40) |
| **Kulmambetova** | 2014 | Kazakhstan | Kazakh | PCR and direct sequencing | *IL1-RN* 2/2 with concomitant *H. pylori* infection increases risk of gastritis | (75) |
| **Kumar** | 2009 | Indian | India | PCR | *IL-1RN* 2/L increases the risk of gastric cancer in *H. pylori* infected patients. *IL-1RN* 2/2 increases the risk of gastric cancer in *H. pylori* uninfected patients. | (76) |
| **Amine** | 2016 | Algeria | Algerian | PCR | *IL1-RN* 2/L with concomitant *H. pylori* infection increases the risk of chronic atrophy gastritis. | (57) |
| **Leea** | 2003 | Korea | Korean | PCR | No association between *IL-1RN* polymorphism and gastric cancer or duodenal ulcer. | (41) |
| **Santos** | 2012 | Brazil | Brazilian | PCR | No association between gastric cancer and *IL-1RN* polymorphism | (77) |
| **Neda** | 2017 | Iran | Iranian | PCR | *IL1-RN* 2/L increases the risk of peptic ulcer | (30) |
| **Rejane** | 2013 | Brazil | Brazilian | PCR | *IL1-RN* 2/2 increases risk of gastric cancer while *IL1-RN* 2/L decreases the risk of gastric cancer *IL-1RN* 2/2 protects against *H. pylori* and peptic ulcer | (34) |
| **Andrew** | 2012 | Kenya | African | PCR-RFLP | *IL1-RN* 2/2 and *IL1-RN* | (70) |
|  |  |  |  |  | L/L increase risk of *H. pylori* infection<br>*IL-IRN* L/L are more frequent in *H. pylori* associated gastric pathologies |  |
| **Bang-Shun** | 2011 | China | Chinese | PCR | *IL1-RN* 2/L with concomitant *H. pylori* infection increases the risk of gastric cancer | (55) |
| **Mansour** | 2006 | Oman | Omani Arab | PCR | *IL1-RN* 2/L with concomitant *H. pylori* infection increases the risk of gastric cancer | (56) |
| **El-Omar** | 2000 | Scotland and Poland | Scottish Polish Caucasian | PCR | *IL1-RN* 2/2 with concomitant *H. pylori* infection increases the risk of gastric cancer | (31) |

*H. pylori* is an extremely common and highly genetically diverse bacteria that are able to alter the host physiology and immune response allowing it to persist for the life of the host. Eradication of *H. pylori* could be a double-edged sword, reducing the incidence of gastric pathologies and gastric cancer while increasing the risk of esophageal diseases and allergies. Many bacterial and Host genotypes act in concert to determine which individuals are at the highest risk of disease. The findings of our study indicate that the *IL-1RN* L/2 genotype is significantly associated with the risk of gastric cancer thus intervention (i.e., eradication treatment) aimed at patients with this genotype has the potential to reduce the risk of gastric cancer substantially.

Notwithstanding the limitations of the present study which include the relatively small number of participants, cross-sectional design, depending on *16S rRNA* for *H. pylori* investigation which did not exclude the previous infection of individual and the investigation has been done in only one gene (*IL-1RN)* without invoking other carcinogenic factors, our findings show that the host genetic can be a useful tool for identifying high-risk individuals among dyspeptic patients; and also underscore the role played by host genetics in gastric carcinogenesis. Further national studies with a large sample size are needed to determine the effect of host- and *H. pylori-* related genotypes on the development of gastric cancer.

## In conclusion

Independently carriage of *IL-1RN* *2 allele contributes significantly to the risk of gastric cancer in the Sudanese population. And this risk is further increased with concomitant *H. pylori* infection. Notwithstanding the relatively small sample size of the study population, our findings show that the host genetic can be a useful tool for identifying high-risk individuals among dyspeptic patients; and also underscore the role played by host genetics in gastric carcinogenesis.

## Data Availability

The data regarding IL-1RN genotypes and alleles distributions among participants that used to support the findings of this study are included within the supplementary information file.

## Acknowledgment

We are very thankful to patients who participated in this study and to the staff of the gastroscopic unit in Ibin Sina specialized hospital, Modern Medical Centre, Al-Shorta hospital and Al Faisal Specialized Hospital.

## Conflict of interest

The authors declare that there are no conflicts of interest.

## Funding Statement

The authors received no specific funding for this work.

## Notes

### Competing Interest Statement

The authors have declared no competing interest.

